# Mendelian randomization integrating GWAS and eQTL data revealed genes pleiotropically associated with major depressive disorder

**DOI:** 10.1101/2020.10.25.20219188

**Authors:** Huarong Yang, Di Liu, Chuntao Zhao, Bowen Feng, Wenjin Lu, Xiaohan Yang, Minglu Xu, Weizhu Zhou, Huiquan Jing, Jingyun Yang

## Abstract

**Objectives:** To prioritize genes that are pleiotropically or potentially causally associated with the risk of MDD.

**Methods:** We applied the summary data-based Mendelian randomization (SMR) method integrating GWAS and expression quantitative trait loci (eQTL) data in 13 brain regions to identify genes that were pleiotropically associated with the risk of MDD. In addition, we repeated the analysis by using the meta-analyzed version of the eQTL summary data in the brain (brain-eMeta).

**Results:** We identified multiple significant genes across different brain regions that may be involved in the pathogenesis of MDD. The prime-specific gene *BTN3A2* (corresponding probe: ENSG00000186470.9) was the top hit showing pleotropic association with MDD in 9 of the 13 brain regions and in brain-eMeta, after correction for multiple testing. Many of the identified genes are located in the human major histocompatibility complex (MHC) region on chromosome 6 and are mainly involved in immune response.

**Conclusions:** Our SMR analysis revealed that multiple genes showed pleiotropic association with MDD across the brain regions. These findings provide important leads to a better understanding of the mechanism of MDD, and reveals potential therapeutic targets for the prevention and effective treatment of MDD.

## Introduction

Major depressive disorder (MDD), also known as clinical depression, is a significant medical condition impacting individual’s mood, behavior as well as appetite and sleep, even thoughts of suicide [1]. MDD is a leading cause of disability and morbidity worldwide [2], with an estimated lifetime prevalence of around 15% [3]. MDD is a complex multi-factorial disorder, with contributions from both genetic and environmental factors [4]. However, the exact etiology of MDD remains to be unclear, and there is pressing urgency to further explore the pathological mechanisms underlying MDD to facilitate the design and implementation of efficient prevention strategies or novel treatments.

Previous twin studies found the heritability of MDD to be approximately 30%– 40% [5,6]. Although genome-wide association studies (GWAS) have been successful in identifying genetic variants associated with MDD [7-11], biological interpretation of the findings remain largely unclear. It is likely that these genetic loci exert their effect on MDD via gene expression. Therefore, it is important to explore the relationship between genetic variation and gene expression to better understand the regulatory pathways underlying the pathogenesis of MDD.

Mendelian randomization (MR) is a popular method for exploring potentially causal association between an exposure (e.g., gene expression) and an outcome (e.g., MDD susceptibility) by using genetic variants as the instrumental variables (IVs) [12]. Compared with traditional statistical methods used in the association studies, MR reduces confounding and reverse causation [13,14], and has been successful in identifying gene expressions or DNA methylation loci that are pleiotropically or potentially causally associated with various phenotypes, such as cardiovascular diseases, inflammatory bowel disease, and educational attainment [15-19].

In this study, we adopted the summary data-based MR (SMR) method integrating summarized GWAS data for MDD and expression quantitative trait loci (eQTL) data in the brain to prioritize genes that are pleiotropically or potentially causally associated with the risk of MDD. Our analysis identified a primate specific gene *BTN3A2* as a novel MDD risk gene of MDD.

## Methods

### Data sources

#### eQTL data

In the SMR analysis, cis-eQTL genetic variants were used as the IVs for gene expression. We performed SMR analysis for different regions in the brain. We used the Version 7 release of the eQTL summarized data from the Genotype Tissue Expression [20] project, which included 13 different regions: amygdala, anterior cingulate cortex, caudate nucleus, cerebellar hemisphere, cerebellum, cortex, frontal cortex, hippocampus, hypothalamus, nucleus accumbens, putamen, spinal cord and substantia nigra [20]. In addition, we repeated the analysis by using the meta-analyzed version of the eQTL summary data (named brain-eMeta hereafter), which included results from GTEx data of brain tissues [20], the Common Mind Consortium [21], and the Religious Orders Study and the Rush Memory and Aging Project [22]. Results from these three studies were meta-analyzed using the MeCS method (meta-analysis of cis-eQTL in correlated samples) to increase the power of detecting brain eQTLs [23]. Only SNPs within 1 Mb distance from each individual probe are available. The eQTL data can be downloaded at https://cnsgenomics.com/data/SMR/#eQTLsummarydata.

#### GWAS data for MDD

The GWAS summarized data for MDD were provided by the Psychiatric Genomics Consortium [10]. The results were based on three large genome-wide association studies [8,9,11], including a total of 807,553 individuals (246,363 cases and 561,190 controls, after excluding overlapping samples) and 8,098,588 genetic variants. The GWAS summarized data can be downloaded at https://www.med.unc.edu/pgc/download-results/mdd/.

### Statistical and bioinformatics analyses

MR was carried out considering cis-eQTL as the IVs, gene expression as the exposure, and MDD as the outcome. MR analysis was performed using the method as implemented in the software SMR. Detailed information regarding the SMR method have been described previously [16]. Briefly, SMR uses the principles of MR integrating GWAS and eQTL summary statistics to test for pleotropic association between gene expression and MDD due to a shared and potentially causal variant at a locus. The heterogeneity in dependent instruments (HEIDI) test was done to explore the existence of linkage in the observed association. Rejection of the null hypothesis (i.e., *P*_HEIDI_<0.05) indicates that the observed association might be due to two distinct genetic variants in high linkage disequilibrium with each other. We adopted the default settings in SMR (e.g., *P*_eQTL_ <5 × 10^−8^, minor allele frequency [MAF] > 0.01, excluding SNPs in very strong linkage disequilibrium [LD, r^2^ > 0.9] with the top associated eQTL, and removing SNPs in low LD or not in LD [r^2^ <0.05] with the top associated eQTL), and used false discovery rate (FDR) to adjust for multiple testing.

Annotations of the transcripts were based on the Affymetrix exon array S1.0 platforms. To functionally annotate putative transcripts, we conducted functional enrichment analysis using the functional annotation tool “Metascape” for the significantly tagged genes in different brain regions and in brain-eMeta. Gene symbols corresponding to putative genes (*P*<0.05) were used as the input of the gene ontology (GO) and Kyoto Encyclopedia of Genes and Genomes (KEGG) enrichment analysis.

Data cleaning and statistical/bioinformatical analysis was performed using R version 4.0.2 (https://www.r-project.org/), PLINK 1.9 (https://www.cog-genomics.org/plink/1.9/) and SMR (https://cnsgenomics.com/software/smr/).

## Results

### Basic information of the summarized data

The number of participants used for generating the eQTL data varied across the brain regions, ranging from 114 to 209, so did the number of eligible probes involved in the final SMR analysis, ranging from 814 to 2,786. The brain-eMeta analysis involved more subjects (n=1,194) and more probes (n=7,421). The GWAS meta-analysis data involved roughly 800,000 subjects. The detailed information was shown in **Table 1**.

**Table 1.** Basic information of the eQTL and GWAS data.

| **Data source** | **Total participants or cases/controls** | **Number of genetic variants or probes** |
| --- | --- | --- |
| **eQTL data** |  |  |
| **Amygdala** | 129 | 779 |
| **Anterior cingulate cortex** | 147 | 1379 |
| **Caudate** | 194 | 2089 |
| **Cerebellar hemisphere** | 175 | 2615 |
| **Cerebellum** | 209 | 3765 |
| **Cortex** | 205 | 2314 |
| **Frontal cortex** | 175 | 1722 |
| **Hippocampus** | 165 | 1108 |
| **Hypothalamus** | 170 | 1170 |
| **Nucleus accumbens** | 202 | 1785 |
| **Putamen** | 170 | 1449 |
| **Spinal cord** | 126 | 915 |
| **Substantia nigra** | 114 | 661 |
| **Brain-eMeta** | 1,194 | 7,421 |
| **GWAS-Meta** | 246,363/561,190 | 8,098,588 |
| **23andme_307k** | 75,607/231,747 |  |
| **UK Biobank** | 127,552/233763 |  |
| **PGC_139k** | 43,204/95,680 |  |

GWAS: genome-wide association studies; QTL, quantitative trait loci; PGC, Psychiatric Genomics Consortium

### SMR analysis in the 13 brain regions

Out of the 13 brain regions, the human major histocompatibility complex (MHC) gene *BTN3A2* (ENSG00000186470.9) was the top hit showing pleotropic association with MDD in 9 regions, after correction for multiple testing. Each of the other two genes, *RPL31P12* (ENSG00000227207.2) and *RP1-265C24*.*5* (ENSG00000219392.1), was the top gene pleiotropically associated with MDD in two brain regions (**Figure S1**).

Specifically, for *BTN3A2*, the most significant associations with MDD were detected in two brain regions: caudate nucleus and spinal cord (β [SE]=0.043 [0.008], *P*=7.76×10^−8^; β [SE]=0.042 [0.008], *P*=1.72×10^−7^; **Figure 1**). It also showed significant pleiotropic association with MDD in the four brain regions where it was not the top gene (**Table S1**). *RPL31P12* showed the most significantly pleiotropic association with MDD in cerebellar hemisphere and cerebellum (β [SE]=-0.037 [0.006], *P*=7.53×10^−11^; β [SE]=-0.033 [0.005], *P*=1.34×10^−12^, respectively; **Figure 2**). *RP1-265C24*.*5* showed significant pleiotropic association in cortex and nucleus accumbens (β [SE]=0.036 [0.007], *P*=6.13×10^−08^; β [SE]=0.036 [0.006], *P*=1.63×10^−08^, respectively; **Figure 3**).

**Figure 1.**
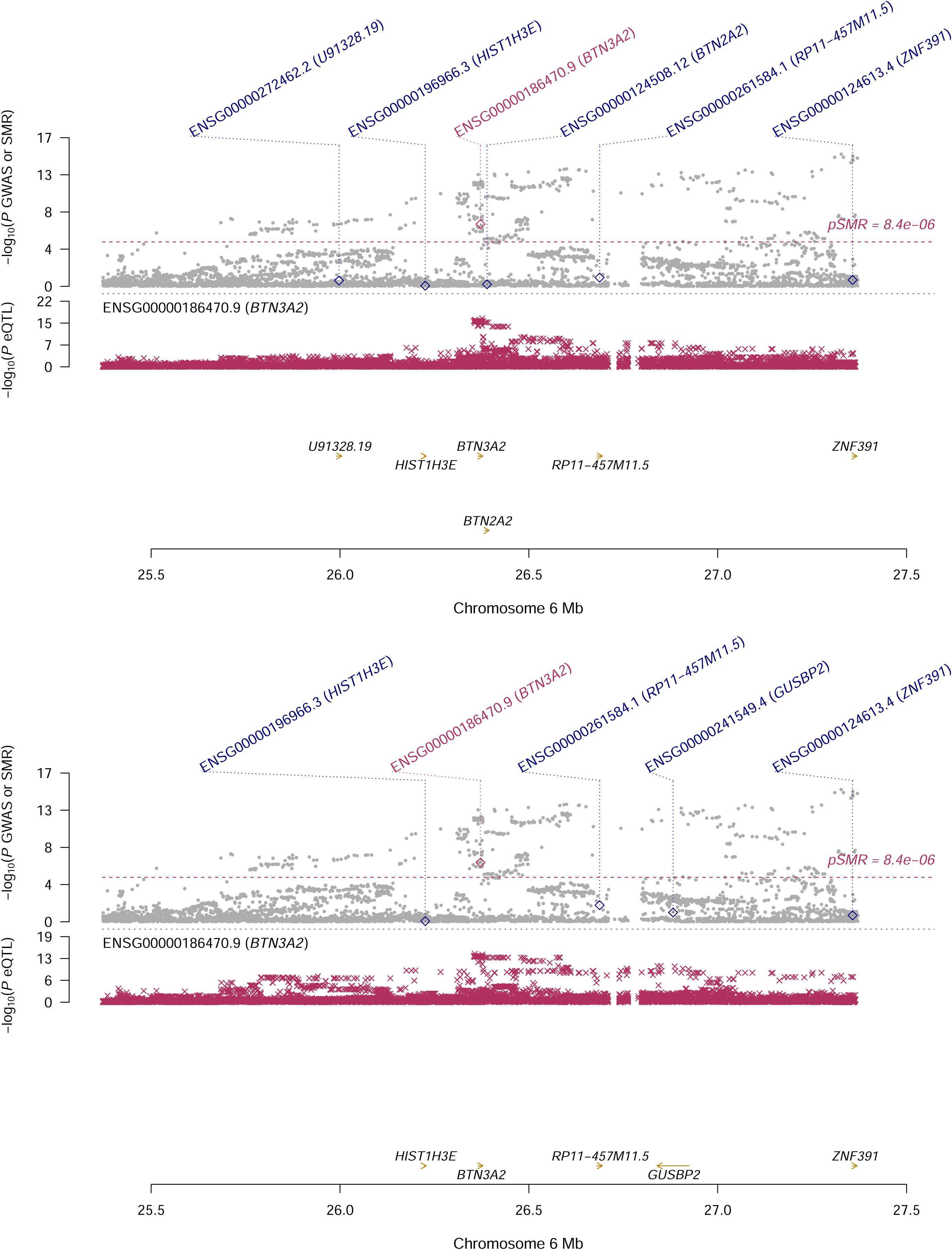
Prioritizing genes around *BTN3A2* in association with MDD. A) Caudate nucleus; B) Spinal cord Top plot, grey dots represent the -log_10_(*P* values) for SNPs from the GWAS of MDD, and rhombuses represent the -log_10_(*P* values) for probes from the SMR test with solid rhombuses indicating that the probes pass HEIDI test and hollow rhombuses indicating that the probes do not pass the HEIDI test. Middle plot, eQTL results for the probe ENSG000001864770.9 tagging *BTN3A2*. Bottom plot, location of genes tagged by the probe. Highlighted in maroon indicates probes that pass the SMR threshold. GWAS, genome-wide association study; MDD, major depressive disorder; SMR, summary data-based Mendelian randomization; HEIDI, heterogeneity in dependent instruments; eQTL, expression quantitative trait loci

**Figure 2.**
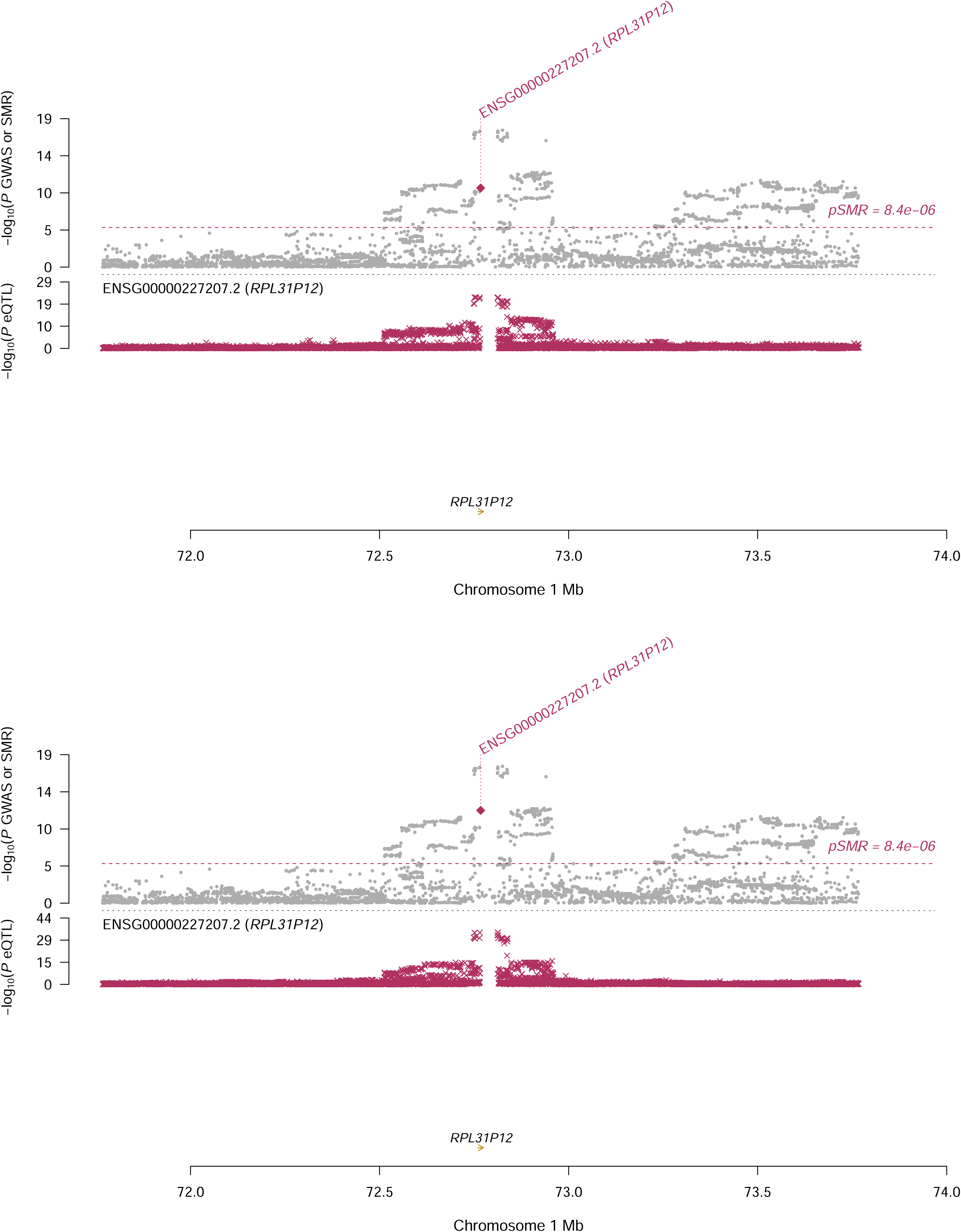
Prioritizing genes around *RPL31P12* in association with MDD. A) Cerebellar hemisphere; B) Cerebellum Top plot, grey dots represent the -log_10_(*P* values) for SNPs from the GWAS of MDD, and rhombuses represent the -log_10_(*P* values) for probes from the SMR test with solid rhombuses indicating that the probes pass HEIDI test and hollow rhombuses indicating that the probes do not pass the HEIDI test. Middle plot, eQTL results for the probe ENSG00000227207.2 tagging *RPL31P12*. Bottom plot, location of genes tagged by the probe. Highlighted in maroon indicates probes that pass the SMR threshold. GWAS, genome-wide association studies; MDD, major depressive disorder; SMR, summary data-based Mendelian randomization; HEIDI, heterogeneity in dependent instruments; eQTL, expression quantitative trait loci

**Figure 3.**
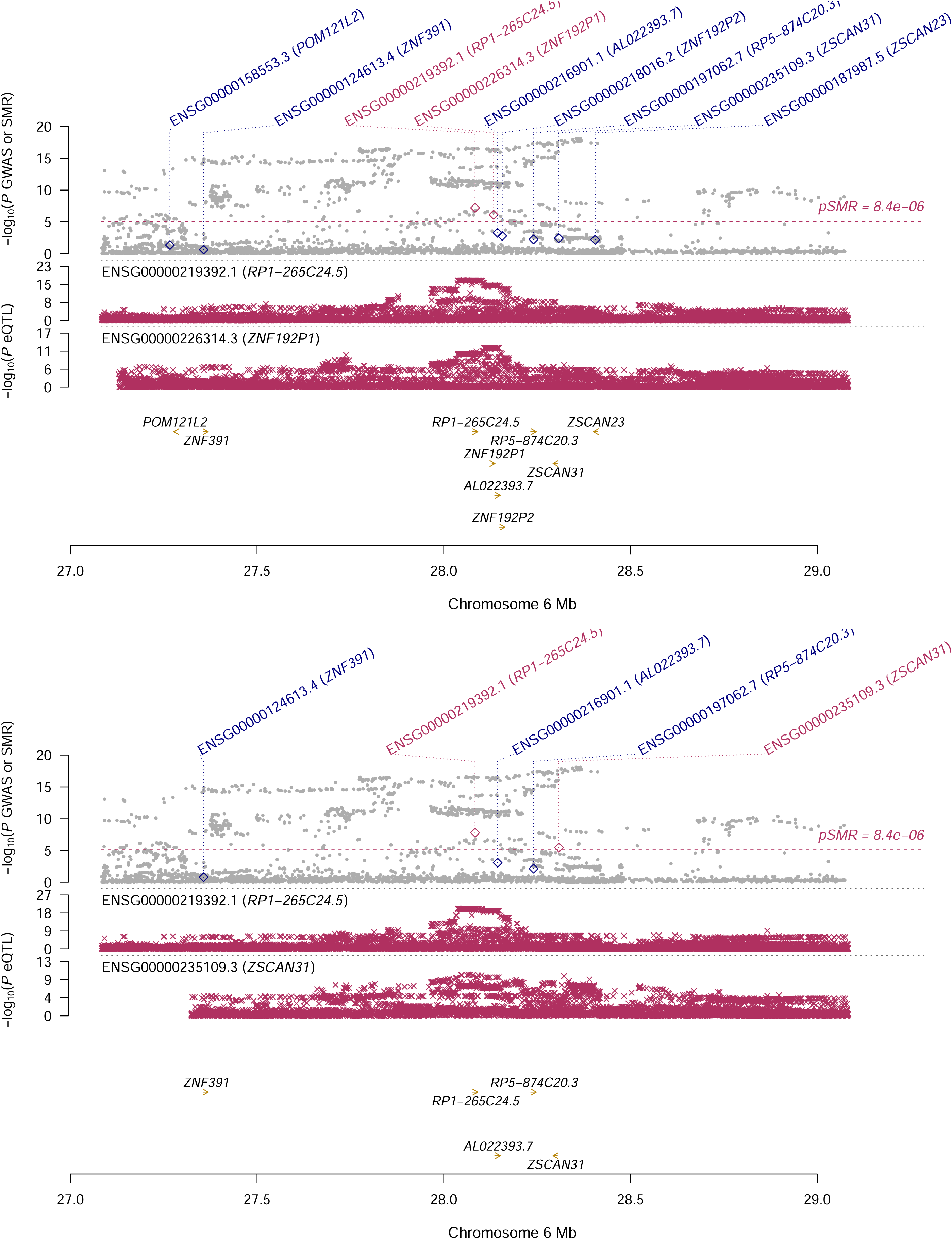
Prioritizing genes around *RP1-265C24.5* in association with MDD. A) Cortex; B) Nucleus accumbens Top plot, grey dots represent the -log_10_(*P* values) for SNPs from the GWAS of MDD, and rhombuses represent the -log_10_(*P* values) for probes from the SMR test with solid rhombuses indicating that the probes pass HEIDI test and hollow rhombuses indicating that the probes do not pass the HEIDI test. Middle plot, eQTL results for the probe ENSG00000219392.1 tagging *RP1-265C24.5*. Bottom plot, location of genes tagged by the probe. Highlighted in maroon indicates probes that pass the SMR threshold. GWAS, genome-wide association studies; MDD, major depressive disorder; SMR, summary data-based Mendelian randomization; HEIDI, heterogeneity in dependent instruments; eQTL, expression quantitative trait loci

The complement gene *C4A* (ENSG00000244731.3) was significantly associated with MDD in 7 different brain regions, after correction for multiple testing (**Table S1**). Of note, both *BTN3A2, C4A* and *RP1-265C24.5* are on chromosome 6 while *RPL31P12* is on chromosome 1. Two brain regions, cerebellar hemisphere and cerebellum, have a relatively large number of significant genes (21 genes and 30 genes, respectively; **Table 2**).

**Table 2.** Summary of the SMR analysis across the 13 brain regions.

| Regions | N_gene | Gene_ID | Gene | CHR | top_SNP | P<sub>SMR</sub> | Q_value |
| --- | --- | --- | --- | --- | --- | --- | --- |
| **Amygdala** | 5 | ENSG00000186470.9 | BTN3A2 | 6 | rs9393703 | $3.08 \times 10^{-07}$ | $2.40 \times 10^{-04}$ |
| **Anterior cingulate cortex** | 5 | ENSG00000186470.9 | BTN3A2 | 6 | rs28551159 | $2.46 \times 10^{-07}$ | $3.40 \times 10^{-04}$ |
| **Caudate** | 7 | ENSG00000186470.9 | BTN3A2 | 6 | rs9379853 | $7.76 \times 10^{-08}$ | $1.62 \times 10^{-04}$ |
| **Cerebellar hemisphere** | 21 | ENSG00000227207.2 | RPL31P12 | 1 | rs1460943 | $7.53 \times 10^{-11}$ | $1.97 \times 10^{-07}$ |
| **Cerebellum** | 30 | ENSG00000227207.2 | RPL31P12 | 1 | rs1460943 | $1.34 \times 10^{-12}$ | $5.03 \times 10^{-09}$ |
| **Cortex** | 7 | ENSG00000219392.1 | RP1-265C24.5 | 6 | rs2295594 | $6.13 \times 10^{-08}$ | $1.42 \times 10^{-04}$ |
| **Frontal cortex** | 4 | ENSG00000186470.9 | BTN3A2 | 6 | rs9379853 | $2.20 \times 10^{-07}$ | $3.78 \times 10^{-04}$ |
| **Hippocampus** | 3 | ENSG00000186470.9 | BTN3A2 | 6 | rs9393703 | $5.32 \times 10^{-07}$ | $5.90 \times 10^{-04}$ |
| **Hypothalamus** | 3 | ENSG00000186470.9 | BTN3A2 | 6 | rs72841536 | $5.34 \times 10^{-07}$ | $6.25 \times 10^{-04}$ |
| **Nucleus accumbens** | 3 | ENSG00000219392.1 | RP1-265C24.5 | 6 | rs4713135 | $1.63 \times 10^{-08}$ | $2.91 \times 10^{-05}$ |
| **Putamen** | 2 | ENSG00000186470.9 | BTN3A2 | 6 | rs9379853 | $3.41 \times 10^{-07}$ | $4.95 \times 10^{-04}$ |
| **Spinal cord** | 2 | ENSG00000186470.9 | BTN3A2 | 6 | rs71557332 | $1.72 \times 10^{-07}$ | $1.58 \times 10^{-04}$ |
| **Substantia nigra** | 1 | ENSG00000186470.9 | BTN3A2 | 6 | rs28551159 | $4.42 \times 10^{-06}$ | $2.92 \times 10^{-03}$ |

Number of genes means the number of statistically significant genes in each region after correction for multiple testing using false discovery rate (Q value < 0.05); top probe and gene is the corresponding gene; top SNP is the top associated cis-eQTL in the eQTL analysis; P<sub>SMR</sub> is the P value for SMR analysis
CHR, chromosome; SNP, single-nucleotide polymorphism; SMR, summary data-based Mendelian randomization; QTL, quantitative trait loci

GO enrichment analysis of biological process and molecular function showed that the significant genes across the different brain regions were involved in four GO terms, including negative regulation of endopeptidase activity (GO:0010951), adaptive immune response (GO:0002250), platelet degranulation (GO:0002576) and negative regulation of defense response (GO:0031348; **Figure S2A**). Concept network analysis of the identified genes revealed multiple domains related with immune response (**Figure S2B**). More information could be found in **Table S2**.

### SMR analysis in the brain-eMeta

Using brain-eMeta eQTL data, we found 75 genes that showed pleiotropic association with MDD, after correction for multiple testing. Specifically, we identified *BTN3A2* (ENSG00000186470) that showed the most significantly pleiotropic association with MDD (β [SE]=0.027 [0.004], *P*=3.44×10^−12^; **Table S3**), followed by *RPL31P12* (ENSG00000227207, β [SE]=-0.039 [0.006], *P*=3.43×10^−11^). We found that *C4A* and *RP1-265C24.5* also showed significant pleiotropic association with MDD (β [SE]=0.031 [0.005], *P*=1.58×10^−8^ and β [SE]=0.047 [0.008], *P*=2.11×10^−09^, respectively).

GO enrichment analysis of biological process and molecular function showed that the significant genes in brain-eMeta were involved in eight GO terms, including allograft rejection (ko05330), butyrophilin (BTN) family interactions (R-HSA-8851680), platelet degranulation (GO:0002576), immunoregulatory interactions between a lymphoid and a non-lymphoid cell (R-HSA-198933), nuclear chromosome segregation (GO:0098813), telomere maintenance (GO:0000723), organelle localization by membrane tethering (GO:0140056) and lipid transport (GO:0006869; **Figure S3A**). Similar to the findings for the different brain regions, concept network analysis in brain-eMeta also revealed multiple domains related with immune response (**Figure S3B**). More information could be found in **Table S4**.

## Discussion

In this study, we integrated GWAS and eQTL data in the MR analysis to explore putative genes that showed pleiotropic/potentially causal association with MDD susceptibility. Across the different brain regions, we identified multiple significant genes that may be involved in the pathogenesis of MDD. The identified genes were mainly involved in immune response. Our findings provided important leads to a better understanding of the mechanisms underlying MDD and revealed potential therapeutic targets for the effective treatment of MDD.

Several of the identified genes in our study, such as *BTN3A2, BTN3A3, PRSS16, HLA-C, C4A* and *HLA-DMA*, are located in or around the human major histocompatibility complex (MHC) region on chromosome 6. MHC represents the most complex genomic region due to its unintelligible linkage disequilibrium [24]. Many genes in or around MHC play an important role in immune response and immune regulation, and are involved in a variety of inflammatory and autoimmune diseases [25-29]. The MHC regions can be roughly divided into three classes that are functionally distinct, with class I and II regions containing highly polymorphic human leukocyte antigen (HLA) genes associated with autoimmune disease risk [30,31] and class III region containing complement component 4 regions associated with schizophrenia risk [32]. Recent GWASs identified a number of genetic variants in the MHC region associated with depression risk, with the strongest association observed in or near the class I region [9-11].

We found that *BTN3A2* were significantly associated with MDD across the brain regions. *BTN3A2*, which encodes a member of the immunoglobulin superfamily, resides in the juxta-telomeric region (class I) of MHC [33]. The BTN3A2 protein may be involved in adaptive immune response [34]. Previous studies showed that *BTN3A2* was a potential risk gene for Alzheimer’s disease, schizophrenia and intellectual disability [35-37]. A meta-analysis of GWAS found that *BTN3A2* was associated with neuroticism [38], an important risk factor for MDD [39]. Overexpression of *BTN3A2* suppressed the excitatory synaptic activity onto CA1 pyramidal neurons, most likely through the interaction with the presynaptic adhesion molecule neurexins [35,40].

Previous research showed that *BTN3A2* was expressed in multiple cell types in the brain, including astrocyte, neuron, oligodendrocyte, and microglia [41]. These findings, together with ours, demonstrated the important role of *BTN3A2* in the nervous system and highlighted the potential of this gene as a promising target for the prevention and treatment of MDD. A previous GWAS of MDD highlighted the importance of the prefrontal brain regions [10]. In the prefrontal cortex, we found four significant genes, including *BTN3A2, C4A, RP1-265C24.5* and *CYP21A1P*, that were associated with MDD after correction for multiple testing. The gene *C4A* was significant in a total of seven brain regions and in brain-eMeta. *C4A* localizes to the MHC class III region and encodes the acidic form of complement factor 4. In the mouse brain, *C4A* gene is mainly expressed in astrocyte and neurons [42]. *C4A* is involved in the classical complement activation pathway [43] and was reported to be associated with schizophrenia, aging and Alzheimer’s disease [32,44,45]. Moreover, genetic variants in *BTN3A2* and *C4A* were in different LD blocks, suggesting that both genes might be independent risk factors for mental disorders such as schizophrenia and MDD [35].

Both MDD and schizophrenia are mental illnesses contributing substantially to the global disease burden. It was reported that depressed patients had a higher risk of developing psychosis. Moreover, even prior to the emergence of psychotic symptoms, patients with a high risk of schizophrenia had a higher risk for developing depressive symptoms [46]. In consistent with previous findings [47], some of the identified genes showing pleiotropic association with MDD were also associated with schizophrenia, such as *BTN3A2, BTN3A3, PRSS16, HLA-C, C4A* and *HLA-DMA*, indicating a potential overlapped mechanism between schizophrenia and MDD. In addition, compared with a previous study [48], the sample size for GWAS of MDD was much large, therefore increasing the power of SMR analysis, and we performed the analysis in different brain regions.

Our study has some limitations. The number of probes used in our SMR analysis were limited for some brain regions (**Table 1**), and we may have missed some important genes. The HEIDI test was significant for some of the identified genes, indicating the possibility of horizontal pleiotropy (supplementary **Table S1** and **Table S3**), i.e., the identified association might be due to two distinct genetic variants in high linkage disequilibrium with each other. In addition, we only included study participants of European ethnicity, and our findings might not be generalized to other ethnicities. More studies are needed to validate our findings in independent populations. We adopted correction for multiple testing to reduce false positive rate; however, we may have missed important SNPs or genes. Due to a lack of individual eQTL data, we could not quantify the changes in gene expression in subjects with MDD in comparison with the control.

## Conclusion

Our SMR analysis revealed that multiple genes showed pleiotropic association with MDD across the brain regions. More studies are needed to explore the underlying physiological mechanisms in the etiology of MDD.

## Data Availability

All data generated or analyzed during this study are included in this published article and its supplementary information files.

## Acknowledgements

The study was supported by NIH/NIA grants P30AG10161, R01AG15819, R01AG17917, R01AG36042, U01AG61356 and 1RF1AG064312-01. Huiquan Jing’s research was supported by National Key Research and Development Program of China (2018YFC2000400). Di Liu was supported by China Scholarship Council (CSC 201908110339).

The authors confirmed that all authors have reviewed the contents of the article being submitted, approved its contents, and validated the accuracy of the data.

## Author contribution

HJ and JY designed the study. HY, BF and WL analyzed data and performed data interpretation. DL, CZ and JY wrote the initial draft, and BF, WL, XY, MX, WZ and HJ contributed to writing the subsequent versions of the manuscript. All authors reviewed the study findings and read and approved the final version before submission.

## Figure legends

**Figure S1. Distribution of genes show significant pleiotropic association with MDD**.

A) A lobe view of the distribution of significant genes; B) a sagittal view of the distribution of significant genes

Plots were generated using R package cerebroViz. Results for cerebellar hemisphere, cortex, nucleus accumbens and spinal cord were not plotted because they were not covered in the package.

AMY, amygdala; CAU, caudate; CB, cerebellum; CNG, anterior cingulate cortex; FL, frontal cortex; HIP, hippocampus; HTH, hypothalamus; MDD, major depressive disorder; PUT, putamen; SN, substantia nigra.

**Figure S2. Functional enrichment and gene concept network analysis based on the identified genes in different brain regions**.

A) Enriched GO terms based on the identified genes in different brain regions; B) Concept network analysis of the identified genes

GO, gene ontology

**Figure S3. Functional enrichment and gene concept network analysis based on the identified genes in brain-eMeta**.

A) Enriched GO terms based on the identified genes in brain-eMeta; B) Concept network analysis of the identified genes

GO, gene ontology

