## Supplementary figures and images for "Mendelian randomization integrating GWAS and eQTL data revealed genes pleiotropically associated with major depressive disorder"

### Supplemental Figure1

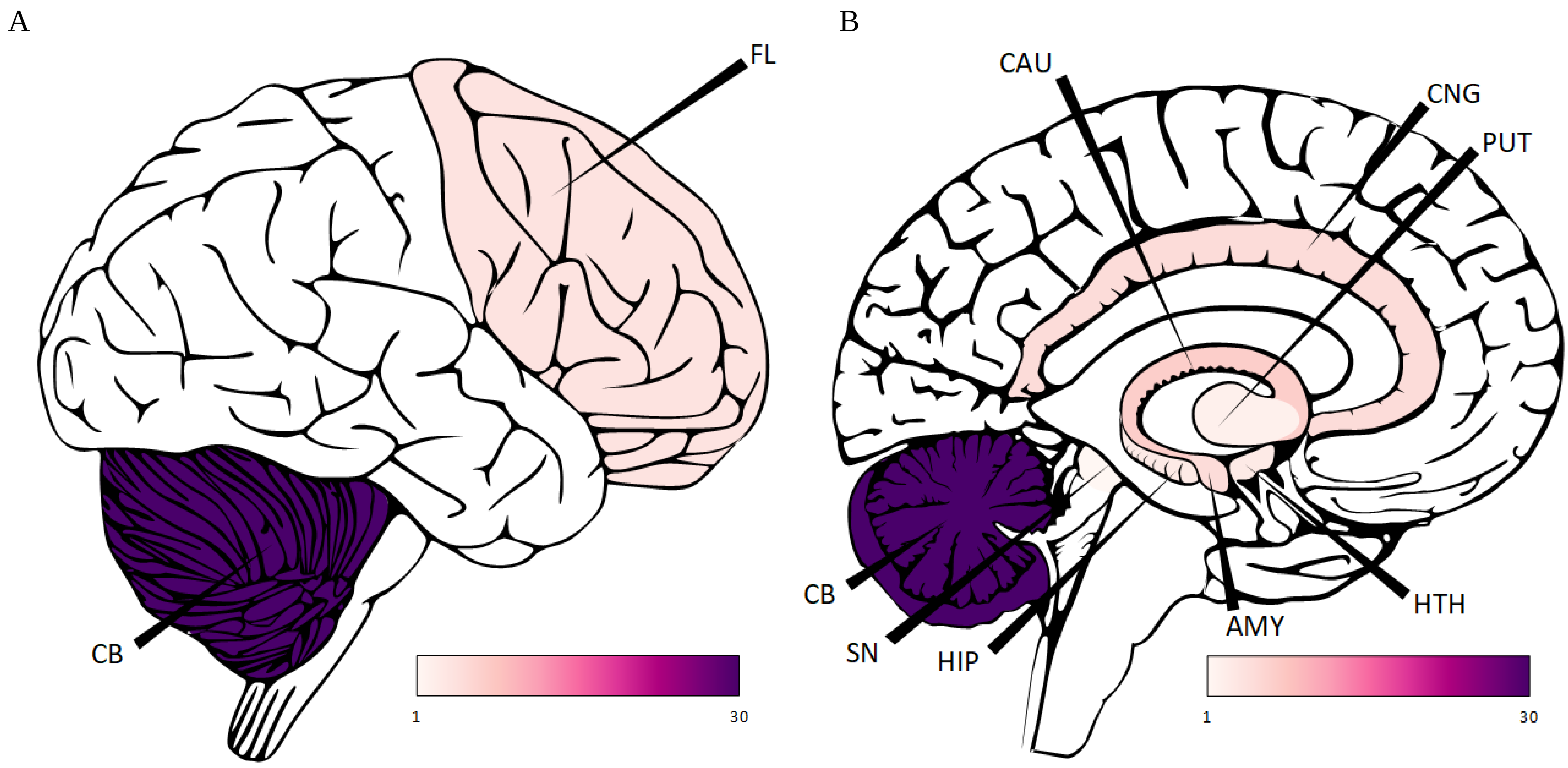

### Supplemental Figure 2

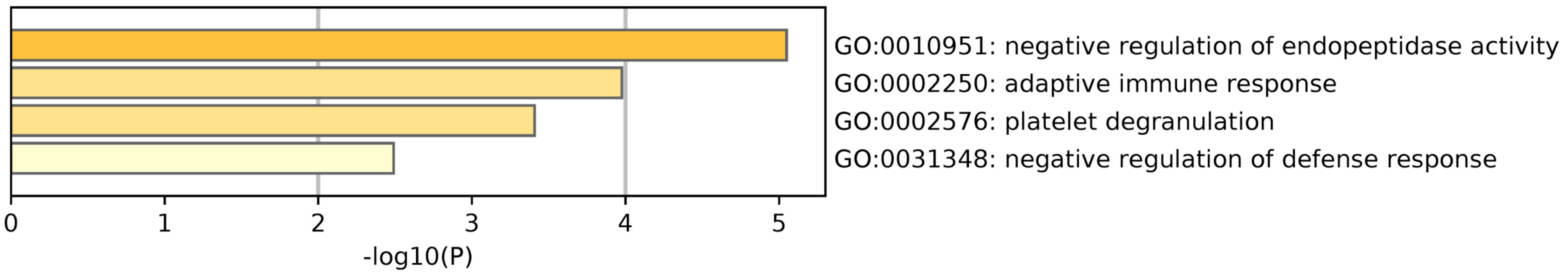

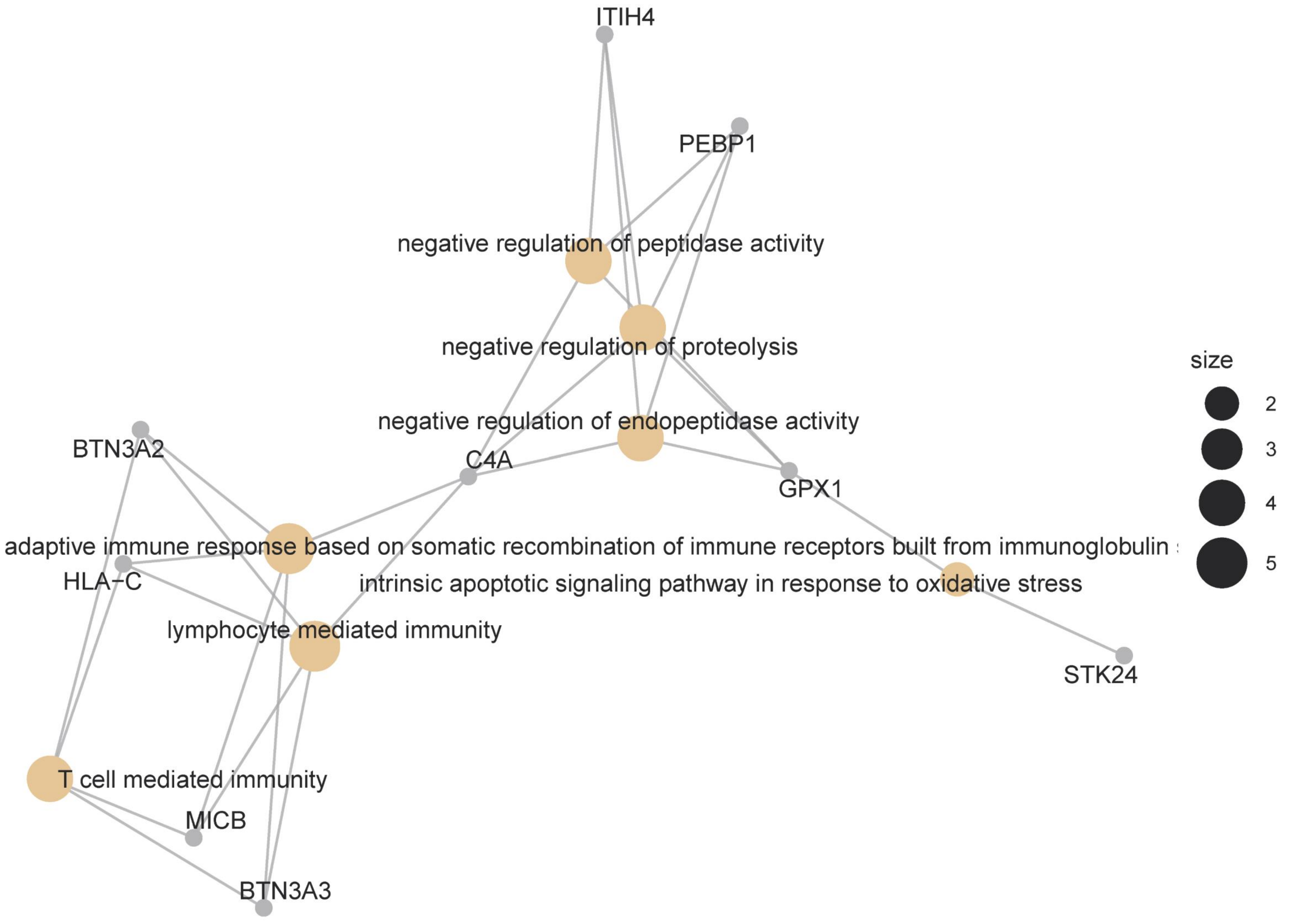

### Supplemental Figure 3

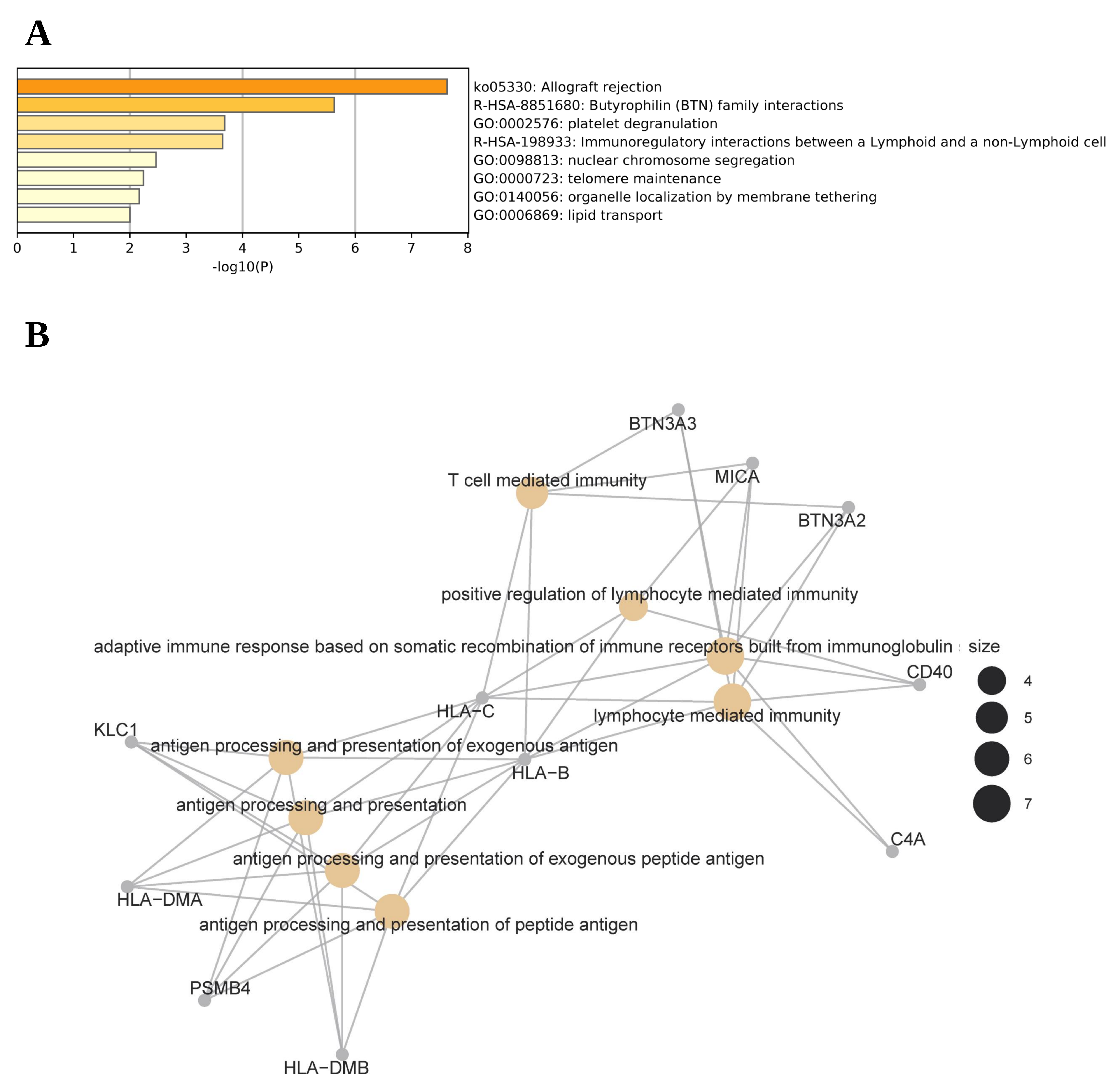
